## Supplementary material for "Actionable absolute risk prediction of atherosclerotic cardiovascular disease: a behavior-management approach based on data from 464,547 UK Biobank participants": S3 Table. Specifications of the python (v3.9.6) libraries and their versions used in this study.

| Library Name | Version |
| --- | --- |
| cycler | 0.10.0 |
| future | 0.18.2 |
| imbalanced-learn | 0.8.0 |
| imblearn | 0.0 |
| joblib | 0.16.0 |
| kiwisolver | 1.3.1 |
| matplotlib | 3.4.2 |
| numpy | 1.21.1 |
| pandas | 1.3.0 |
| Pillow | 8.3.1 |
| pip | 21.1.3 |
| pyparsing | 2.4.7 |
| python-dateutil | 2.8.1 |
| python-dotenv | 0.18.0 |
| pytz | 2020.1 |
| Scikit-learn (1) | 0.24.2 |
| scipy | 1.7.1 |
| seaborn | 0.11.1 |
| setuptools | 57.0.0 |
| six | 1.15.0 |
| threadpoolctl | 2.1.0 |
| xgboost (2,3) | 1.4.2 |

1: Pedregosa F, Varoquaux G, Gramfort A, Michel V, Thirion B, Grisel O, et al. Scikit-learn: Machine Learning in Python. Journal of Machine Learning Research. 2011;12(85):2825–30.

2. Chen T, Guestrin C. XGBoost: A Scalable Tree Boosting System. Proceedings of the 22nd ACM SIGKDD International Conference on Knowledge Discovery and Data Mining. 2016 Aug 13;785–94.

3. XGBoost Documentation — xgboost 1.6.0-dev documentation [Internet]. [cited 2021 Nov 8]. Available from: https://xgboost.readthedocs.io/en/latest/
