## Supplementary material for "Actionable absolute risk prediction of atherosclerotic cardiovascular disease: a behavior-management approach based on data from 464,547 UK Biobank participants": S4 Table. List of utilized open-source methods, best parameters and references.

| Method name and references | Best performing parameters* |
|---|---|
| Extreme Gradient Boosting (XGBoost) (1-3) | subsample: 0.8, n_estimators: 1500, max_depth: 5, learning_rae': 0.01, gamma: 1, colsample_bytree: 0.6 |
| Logistic Regression with L1 regularization and saga solver (4,5) | solver=saga, max_iter=1000, penalty='l1' |
| Random Forest (4,6) | n_estimators=20, criterion='entropy', max_depth=None, min_samples_split=2, min_samples_leaf=1, max_features='auto' |

* Best performing parameters found using Grid Search, default parameters were used if not stated otherwise


1. Friedman JH. Greedy Function Approximation: A Gradient Boosting Machine. The Annals of Statistics. 2001;29(5):1189–232.

2. Chen T, Guestrin C. XGBoost: A Scalable Tree Boosting System. Proceedings of the 22nd ACM SIGKDD International Conference on Knowledge Discovery and Data Mining. 2016 Aug 13;785–94.

3. XGBoost Documentation — xgboost 1.6.0-dev documentation [Internet]. [cited 2021 Nov 8]. Available from: https://xgboost.readthedocs.io/en/latest/

4. Pedregosa F, Varoquaux G, Gramfort A, Michel V, Thirion B, Grisel O, et al. Scikit-learn: Machine Learning in Python. Journal of Machine Learning Research. 2011;12(85):2825–30.

5. McCullagh P, Nelder JA. Generalized linear models. Routledge; 2019.

6. Breiman L. Random Forests. Machine Learning. 2001 Oct 1;45(1):5–32.
